# Human Papillomavirus-Associated Nasopharyngeal Carcinoma: A Systematic Review and Meta-Analysis

**DOI:** 10.1101/2024.09.10.24313140

**Authors:** Brian Y. Zhao, Shun Hirayama, Deborah Goss, Yan Zhao, Daniel L. Faden

## Abstract

**Objective:** Human papillomavirus (HPV) is known to affect head and neck sites beyond the oropharynx, including the nasopharynx. Unlike HPV-associated oropharyngeal squamous cell carcinoma (HPV+OPSCC), HPV-associated nasopharyngeal carcinoma (HPV+NPC) is not well characterized and the true prevalence in non-endemic regions is poorly described. Here, we sought to obtain a global point prevalence of HPV in NPC, stratified by geographic region.

**Data Sources:** EMBASE, OVID Medline, and Web of Science were systematically searched for available evidence on September 21, 2022 for articles published between January 1, 1990 and September 21, 2022.

**Review Methods:** We reviewed the literature for all studies examining NPC and HPV status in adult patients that provided a quantitative HPV prevalence. The study followed the Preferred Reporting Items for Systematic Reviews and Meta-Analyses (PRISMA) guidelines. Main outcome and measures included HPV+NPC prevalence estimates stratified by geographic region, along with other clinical and demographic features.

**Results:** Of the 1567 citations retrieved, 46 studies encompassing 6314 NPC patients were eligible for statistical analysis. The global prevalence of HPV+NPC was 0.18 (95% CI 0.14-0.23). When stratified by geographic region, prevalence was highest in North America (0.25, 95% CI 0.17-0.36), which is a non-endemic region for NPC and also has highest prevalence for HPV+OPSCC. Asia, an endemic area, had the lowest HPV prevalence estimate (0.13, 95% CI 0.08-0.22). HPV 16 (44%) and 18 (33%) were the predominant genotypes in HPV+NPC, dissimilar to HPV+OPSCC.

**Conclusion:** This systematic review and meta-analysis provides a global point prevalence of HPV+NPC stratified by geographic region and suggests that HPV is a significant etiological factor of NPC in North America.

## Introduction

Nasopharyngeal carcinoma (NPC) is classically associated with Epstein-Barr virus (EBV) and has a striking geographic distribution with highest rates in certain parts of Asia^1^. Human papillomavirus (HPV), another oncogenic virus, has a well-established role in oropharyngeal squamous cell carcinoma (HPV+OPSCC), with increasing HPV+OPSCC incidence in North America and western Europe. HPV is known to be an etiologic agent in other head and neck cancer subsites, primarily the nasal tract, including NPC^2–6^. However, the relative rarity of NPC and large variability between geographic regions have led to a heterogeneous collection of studies^7^. Thus, unlike EBV+NPC, characterization of HPV+NPC remains limited. While most NPC cases in endemic regions have been found to be EBV-associated, there is no similar understanding of how HPV+NPC prevalence estimates differ by geographical area and particularly, in geographic regions with a high HPV+OPSCC incidence.

Here, we present a systematic review and meta-analysis of the existing HPV+NPC literature, identifying 6314 NPC cases for inclusion. We aimed to establish a reliable point prevalence of HPV+NPC, both globally and stratified by geographic region, and describe its clinical characteristics. In doing so, we aimed to explore orthogonal hypotheses supporting the involvement of HPV in NPC tumorigenesis such as HPV+NPC prevalence being highest in non-endemic geographic regions with a high HPV+OPSCC prevalence.

## Methods

### Search Strategy

A systematic review was performed following the guidelines of the Preferred Reporting Items for Systematic Reviews and Meta-Analyses (PRISMA)^8^. A search of published studies in Medline via Ovid (1946-), Embase.com (1947-), and Web of Science -All Databases (1900-) was performed on 21 September, 2022 to identify relevant articles. Each search utilized a combination of controlled vocabulary and keywords focused on the terms: human papillomavirus, nasopharyngeal carcinoma, and nasopharynx (**Supplemental Figure 1**). The search was constructed to exclude non-human studies. Filters for a publication year of 1990 or later and written in either English or Japanese (languages spoken by the reviewing authors #1 and #2) were used in the search (**Figure 1**). The search yielded 1567 results before deduplication, and 1163 after.

**Figure 1.**
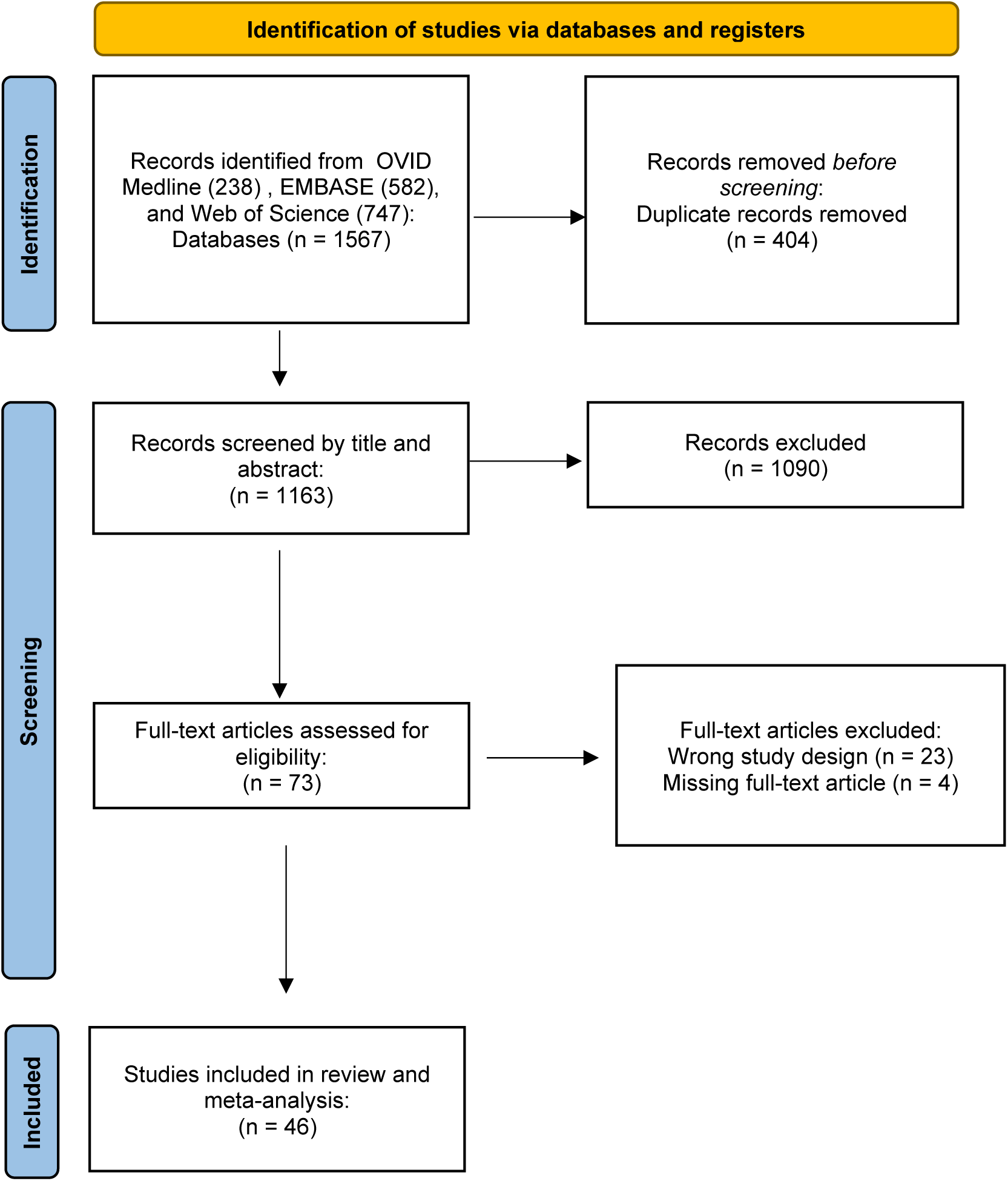
Preferred Reporting Items for Systematic Reviews and Meta-Analyses (PRISMA) flowchart depicting the study selection process.

### Selection Criteria

All articles were compiled and imported into Covidence, for study screening, selection, and data extraction. Titles and abstracts were screened for inclusion by two independent reviewers, with all abstracts, case reports, systematic reviews, and meta-analyses and nonhuman studies being excluded. Studies in English or Japanese from 1990 to September 21, 2022 were included. Any disagreements in the screening process were settled by discussion and consensus between the two authors. All studies examining NPC and HPV status in adult patients that provided a quantitative HPV prevalence were eligible for inclusion. The full texts of the remaining 73 articles were reviewed by two independent reviewers. To avoid double-counting of patients, both reviewers examined each article to determine the tissue bank or database that samples were obtained from and confirm no overlap of sources. 46 articles were ultimately selected for inclusion.

### Data Collection

From the 46 articles selected for inclusion, the following data points were extracted: year of study, country of origin of study patients, sample size, sex and race/ethnicity of study patients, HPV detection assay, HPV genotype, EBV positivity, and World Health Organization (WHO) tumor classification. All eligible studies were used to conduct the final statistical analysis. All studies were screened for duplicate data by comparing authors, timeframe of data collection, and outcomes.

### Statistical Analysis

Statistical analyses were performed using the statistical software R version 4.1.2 (R Foundation). The proportion of HPV+NPC in **each study** were combined to give a pooled HPV prevalence from all eligible studies using random-effects meta-analysis. The pooled estimates were presented together with the associated 95% CIs using forest plots. Meta-regression analyses were implemented in subgroups based on geographic region, race/ethnicity, sex, WHO classification, and HPV detection method. The I2 statistic was used to measure the magnitude of heterogeneity between studies and was categorized according the Cochrane Handbook for Systematic Reviews of Interventions as not important (0%-40%), moderate (30%-60%), substantial (50%-90%), or considerable (75%-100%) heterogeneity^9^. An assessment of publication bias was performed by generating a funnel plot and assessing asymmetry with Egger’s Test.

## Results

### Study characteristics

We identified 46 studies that met criteria for inclusion, containing a total of 6314 NPC patients (**Table 1**)^10–55^. The largest number of studies were from Asia (19) and North America (17). For studies reporting detailed HPV detection methodology, the most common method was polymerase chain reaction (PCR)-based, followed by p16 immunohistochemistry and in situ hybridization (ISH). 37 studies were performed using formalin-fixed-paraffin-embedded (FFPE) tissue samples. Of the studies that reported the HPV genotype, there was a broad spectrum of HPV subtypes detected, including high-risk subtypes such as 16, 18, 31, 33, 35, 39, 45, 52, and 59, low-risk subtypes such as 6 and 11, and unknown-risk subtypes such as 7 and 26. HPV 16 and HPV 18 were the most common genotypes, at 44% and 33% respectively, a striking difference from HPV+OPSCC which is dominated by HPV 16 (**Figure 2**) ^56^.

**Figure 2.**
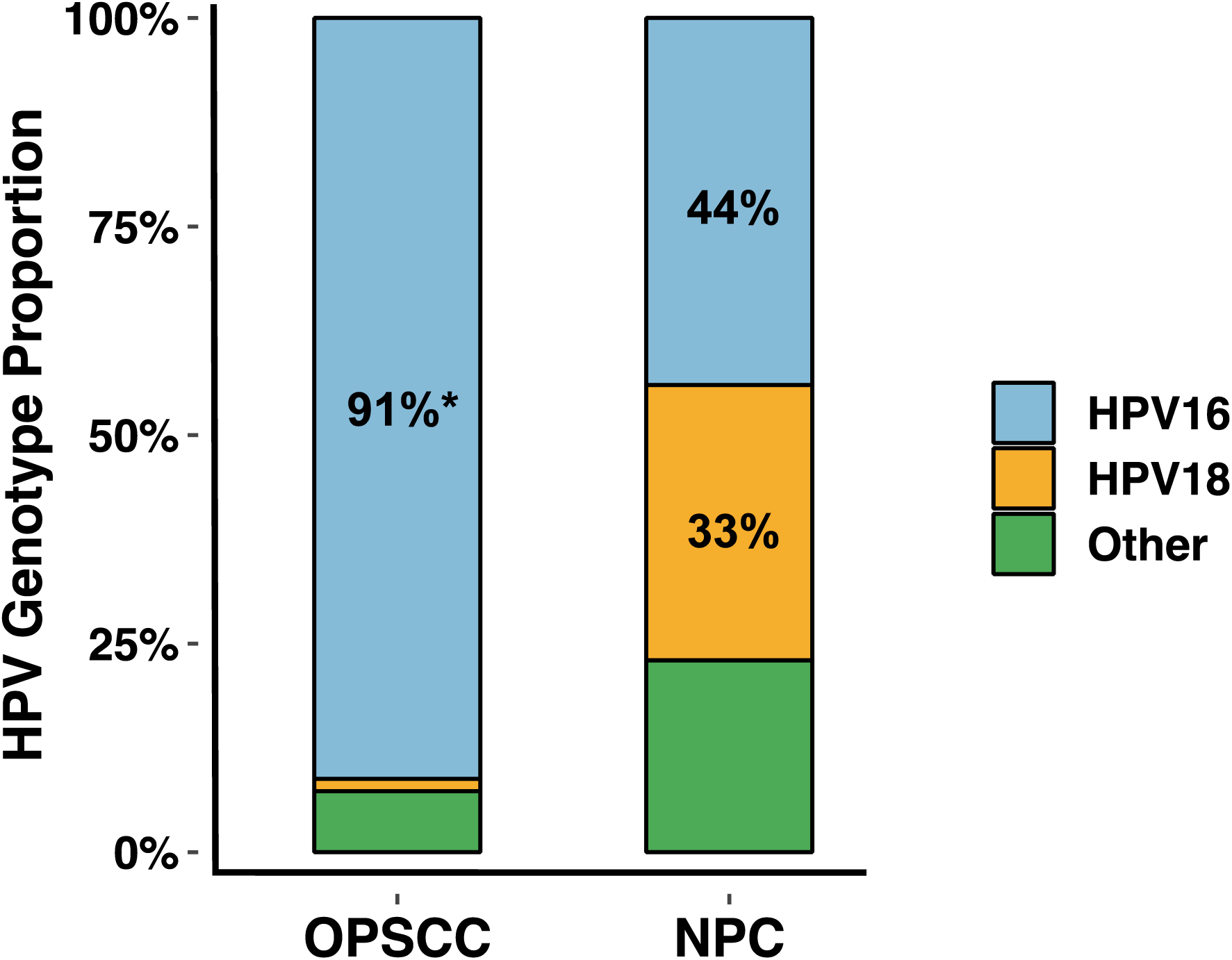
HPV genotype distribution in nasopharyngeal carcinoma and oropharyngeal squamous cell carcinoma. * Source: Ndiaye et al., (2014)

**Table 1.** Overview of studies included in metanalysis (6314 cases total). Key study characteristics are shown including geographic region represented, HPV detection method, and HPV prevalence.

| *First Author* | *Publication Year* | *Geographic Region* | *HPV Detection Assay* | *HPV+/Total* |
| --- | --- | --- | --- | --- |
| Dickens | 1992 | Asia | PCR | 0/16 |
| Tyan | 1993 | Asia | PCR | 14/30 |
| Hording | 1994 | Europe | PCR | 4/38 |
| Giannoudis | 1995 | Europe | PCR | 12/63 |
| Rassekh | 1998 | North America | PCR | 9/17 |
| Kassim | 1998 | Africa | PCR | 5/20 |
| Tung | 1999 | Asia | PCR | 45/88 |
| Punwaney | 1999 | Asia, North America | PCR | 10/30 |
| Lopez-Lizarraga | 2000 | North America | PCR | 13/16 |
| Mirzamani | 2006 | Asia | ISH | 4/20 |
| Maxwell | 2009 | North America | PCR | 4/5 |
| Lo | 2010 | North America | PCR; ISH; p16 | 25/30 |
| Laantri | 2011 | Africa | PCR | 24/70 |
| Huang | 2011 | Asia | PCR; ISH; p16 | 32/89 |
| Deng | 2011 | Asia | PCR | 3/9 |
| Barwad | 2011 | Asia | PCR | 1/20 |
| Singhi | 2012 | North America | ISH; p16 | 4/45 |
| Walline | 2013 | North America | PCR; ISH; p16 | 9/18 |
| Robinson | 2013 | Europe | PCR; ISH; p16 | 11/67 |
| Lin | 2013 | Asia, North America | PCR; ISH; p16 | 10/194 |
| Dogan | 2014 | North America | ISH; p16 | 6/63 |
| Deng | 2014 | Asia | PCR | 6/20 |
| Atighechi | 2014 | Asia | PCR | 9/41 |
| Jiang | 2015 | North America | ISH; p16 | 40/86 |
| Svajdler Jr. | 2016 | Europe | PCR; ISH; p16 | 1/62 |
| Dogan | 2016 | Asia | ISH; p16 | 1/82 |
| Saito | 2017 | Asia | ISH; p16 | 7/43 |
| Kano | 2017 | Asia | PCR; p16 | 2/59 |
| D'Souza | 2017 | North America | ISH; p16 | 13/125 |
| Chang | 2017 | Asia | ISH | 0/56 |
| Asante | 2017 | Africa | PCR | 14/72 |
| Huang | 2018 | Asia | ISH; p16 | 102/1328 |
| Sekee | 2018 | Africa | PCR; p16 | 1/3 |
| Ruuskanen | 2019 | Europe | PCR; ISH; p16 | 21/150 |
| Wotman | 2019 | North America | N/A | 180/517 |
| Wookey | 2019 | North America | N/A | 402/1244 |
| Kofi | 2019 | Africa | PCR | 0/9 |
| Verma | 2020 | North America | PCR; ISH; p16 | 21/343 |
| Simon | 2020 | Europe | PCR; ISH; p16 | 18/98 |
| Bulane | 2020 | Africa | PCR | 1/26 |
| Altekin | 2020 | Asia | ISH | 3/56 |
| Wu | 2022 | North America | ISH; p16 | 12/78 |
| Simon | 2022 | Europe | PCR; ISH | 4/30 |
| Shimizu | 2022 | Asia | p16 | 9/81 |
| Scott-Wittenborn | 2022 | North America | ISH; p16 | 16/127 |
| Huang | 2022 | North America | PCR; p16 | 29/630 |

### HPV Prevalence

Across 46 studies, HPV was reported to be detected in 1154/6314 cases. Using the random-effects model, a global prevalence was estimated at 0.18 (95% CI 0.14-0.23) (**Figure 3**).

**Figure 3.**
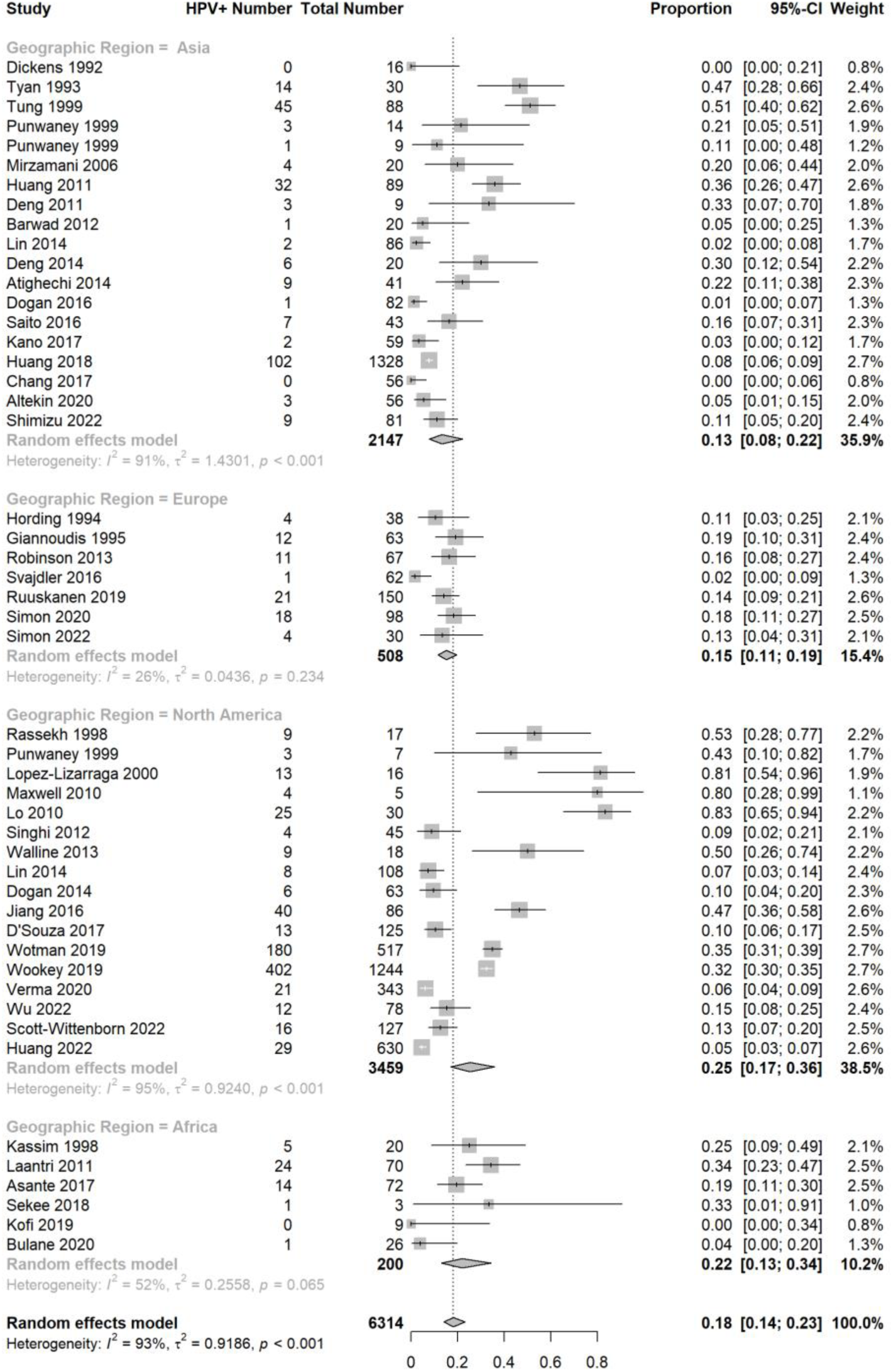
Forest plot of HPV+NPC prevalence estimates using the random-effects model stratified by geographic region, along with a global prevalence estimate.

### Geographic Region

Study geographic region was stratified into four major areas: Africa, Europe, Asia, and North America. Two studies contained cases from multiple geographic areas and were thus split into multiple cohorts, one for each geographic area, for the purposes of the meta-analysis^34,42^. There were no studies from South America or Oceania.

Using the random-effects model, the highest HPV prevalence estimate was found in North America, a non-NPC endemic area, at 0.25 (95% CI 0.17-0.36), while Asia, an endemic area, had the lowest HPV prevalence estimate at 0.13 (95% CI 0.08-0.22) (**Figure 3**). The HPV prevalence estimates in Europe and Africa fell in-between, at 0.15 (95% CI 0.11-0.19) and 0.22 (95% CI 0.13-0.34), respectively. These trends closely mirrored our prior findings in HPV+OPSCC and HPV-associated sinonasal squamous cell carcinoma (HPV+SNSCC) (**Figure 4**). A meta-regression analysis that accounted for the heterogeneity between studies was performed, showing a significant difference between the North American and Asian HPV prevalence estimates (p=0.047). A funnel plot was generated including all studies; Egger’s test showed no statistical evidence for publication bias with an intercept of 0.3939 (95% CI 0.3059-0.4819), with t = 0.6481, df = 47, and p-value of 0.5201 (Supplemental Figure 7).

**Figure 4.**
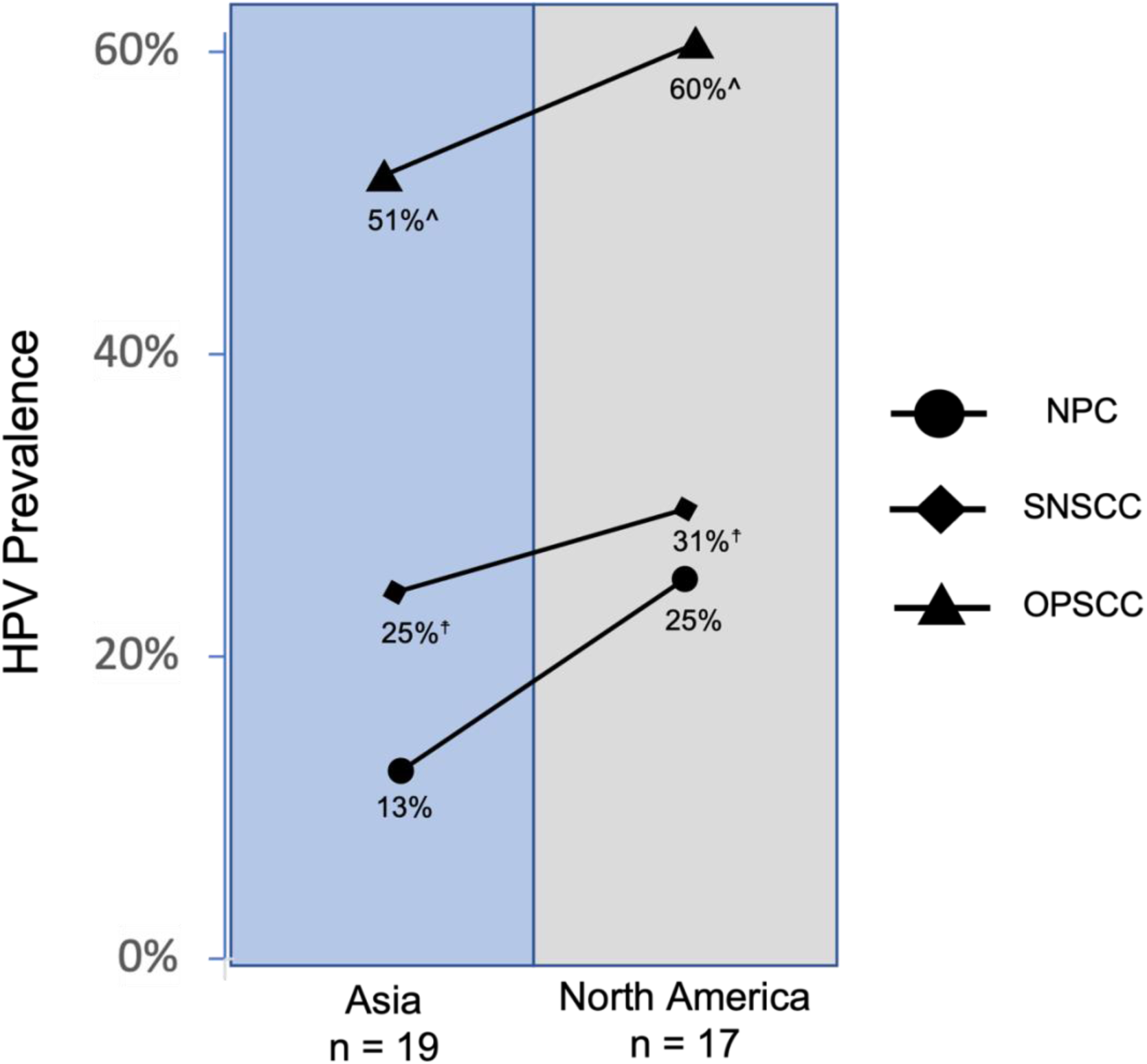
HPV prevalence in NPC, OPSCC and SNSCC in Asian and North America, demonstrating paired prevalence rates across all anatomic sites. ^ Source: Ndiaye et al., (2014). ☨ Source: Pang et al., (2020).

### Race/Ethnicity

A meta-analysis was performed on all North American and UK studies that reported the race/ethnicity of the study patients, with a total of 2767 NPC patients represented. The HPV prevalence estimate for white patients was 0.26 (95% CI 0.16-0.38) while it was 0.16 (95% CI 0.08-0.28) and 0.11 (95% CI 0.06-0.17) for Black and Asian patients, respectively (**Supplemental Figure 2**). There were only 13 Hispanic patients identified in the analysis, which yielded an estimate with a broad 95% confidence interval at 0.42 (95% CI 0.12-0.80). A meta-regression analysis demonstrated a significant difference in prevalence estimates between Asian and white patients (p = 0.008).

### Sex

A meta-analysis was performed on all studies that reported the sex of the study patients, with a total of 2767 NPC patients represented. Of these, 2205 patients were male (79.7%) and 562 were female (20.3%) **(Supplemental Figure 3).** The HPV prevalence estimate for both sexes was found to be identical at 0.17, with similar 95% confidence intervals for males (95% CI 0.10-0.28) and females (95% CI 0.11-0.26). A meta-regression analysis found no significant difference between male and female HPV prevalence estimates (p=0.869).

### WHO Classification

A meta-analysis was performed on all studies that reported the WHO classification of their study samples, with a total of 2359 NPC patients represented. 5.8% of these patients (136/2359) were classified as WHO I, with the rest all classified as WHO II/III. The HPV prevalence estimate in WHO I NPC was 0.39 (95% CI 0.27-0.52), much higher than the estimate of 0.16 (95% CI 0.27-0.52) in WHO II/III patients **(Supplemental Figure 4**). A meta-regression analysis found a significant difference between the HPV prevalence estimates for WHO I and WHO II/III classified tumors (p=0.011).

### HPV Detection Method

Of the 40 studies that detailed HPV detection methods, 19 studies relied solely on PCR-based techniques. Only one study used p16 staining as their sole HPV detection method. All other studies used a mixture of PCR, ISH, and/or p16 staining to determine HPV positivity. Of these, a meta-analysis was performed on the 20 studies that distinguished between p16-only positivity and non-p16-based methodology positivity. Using a random effects model, the HPV prevalence estimate using only p16 was higher at 0.17 (95% CI 0.11-0.26) than when using a non-p16 methodology at 0.14 (95% CI 0.10-0.21) (**Supplemental Figure 5**), with a meta-regression analysis showing no significant difference between the two categories.

### EBV and HPV Co-Infection

20 studies assessed samples for both HPV and EBV status, the latter of which was detected using nucleic acid-based methods. EBV prevalence was found to be 0.65 (95% CI 0.54-0.74) The prevalence of HPV and EBV co-infection was found to be 0.03 (95% CI 0.02-0.07), shown to be significantly lower than that of either virus by itself via a meta-regression analysis (**Supplemental Figure 6**).

## Discussion

We performed a comprehensive meta-analysis of existing studies evaluating HPV in NPC to provide a contemporary understanding of HPV prevalence in NPC, using the largest cohort to date, and spanning four major geographic regions. Using a random-effects model, we estimated the global prevalence of HPV+NPC to be 18%. When further stratified by geography, there was considerable variation, with an intriguing pattern of HPV+NPC prevalence across the geographic regions. In North America and Asia, the two regions containing the most studies and having the most powered analyses, the pattern is in line with our original hypotheses—HPV prevalence is highest in North America (25%), a non-NPC endemic region, and the lowest in Asia (13%), an NPC endemic region^1^. Taken together with the extremely low rate of EBV/HPV co-infection that we observed (3%), HPV is now a significant etiology of NPC in North America.

As we hypothesized, HPV+NPC incidence trends with HPV+OPSCC and HPV+SNSCC incidence, suggesting that the same risk factors that drive the high incidence of HPV+OPSCC in North America are also at play in NPC. Risk factors for HPV+OPSCC are well established. For example, the risk of HPV+OPSCC is most closely associated with the number of oral sex partners, practices that are more common in North America compared to Asia^57,58^. As a site contiguous with the oropharynx, the nasopharynx is assumed to be routinely exposed to HPV-containing saliva. In the oropharynx, HPV has a known tropism for the lymphoid epithelium of the lingual and palatine tonsils; it is thus possible that the lymphoid tissue in the nasopharynx such as adenoids is also hospitable to HPV infection^59^. In fact, a recent study demonstrated that HPV+NPC arises from a more medial anatomic location within the nasopharynx, consistent with a lymphoid origin in the adenoid pad, compared to EBV+NPC, which has classically arisen from the fossa of Rosenmuller, suggesting a different spatial propensity in HPV+NPC^11^. Furthermore, a previous study on HPV+SNSCC found an increased HPV prevalence in subsites with the greatest anatomical susceptibility to oropharyngeal secretion reflux, such as the nasal septum, suggesting that HPV infection and subsequent tumorigenesis in head and neck sites is indeed influenced by the degree of exposure to HPV-infected saliva^4^.

Viral competition is another consideration. Gause’s Law states that when two sympatric non-interbreeding populations compete for the same ecological niche, one displaces the other^60^. In cervical cancer, there has been some evidence of EBV as a cofactor or mediator in HPV-tumorigenesis; however, in head and neck cancer, co-infection has been consistently rare^61–64^. In our analysis, we found that HPV+NPC is significantly more likely to be classified as WHO Type I (39%) than WHO Type II/III (16%), in line with previous studies. In contrast, EBV+NPC is almost always found to be Type WHO II/III, further supporting the hypothesis that EBV-based tumorigenesis is distinct from that of HPV^65^. Thus, in non-endemic areas where EBV+NPC is rare, HPV+NPC prevalence may be high due to less competitive pressure within the cell.

Variation in the genetic predisposition to HPV infection for the population of each geographic region may also contribute to the geographic disparity in HPV+NPC prevalence. Previous studies have shown that Asian men have the lowest incidence of HPV infection while White men have the highest incidence—differences that persist even after adjusting for potential confounding and sexual behavior factors^66,67^. In North American and UK studies, regions with diverse ethnic populations, we found the same trend in HPV+NPC, with the highest incidence in White patients at 26% and the lowest in Asian patients at 11%. Genetic factors involving immune response, including HLA variants that have been associated with European and Asian-ancestry populations, have been shown to influence HPV infection susceptibility^68^. There are also inherent geographic differences among HPV genotypes, with each genotype having its own rate of acquisition and clearance, that may be affecting HPV+NPC distribution.

Beyond geographic prevalence differences, we discovered other notable characteristics of HPV+NPC. In NPC studies with genotyping, HPV 16 (44%) and HPV 18 (33%) emerged as the predominant genotypes. This differs from the oropharynx, where HPV 16 overwhelmingly dominates, contributing to approximately 90% of tumors^69^. A growing body of work has suggested that HPV genotypes are unevenly distributed across head and neck anatomic subsites. Importantly, we have previously demonstrated that non-OPSCC HPV-associated cancers such as HPV+SNCC similarly have significantly lower rates of HPV 16^5,6^. Much remains unknown about the tumorigenic and prognostic differences among the high-risk HPV genotypes, and the significance of HPV 18, and other non-16 genotypes being more common in NPC than other subsites is unclear. In cervical cancer, HPV 18 has been associated with a more aggressive small-cell phenotype; further studies looking at outcomes will be needed to determine if HPV18+NPC also represents a unique clinical entity^70^.

We found no significant difference in HPV+NPC prevalence between sexes, suggesting that HPV+NPC may not have the same male predilection as HPV+OPSCC. In addition, we found that p16 based testing yielded only a slightly higher prevalence estimate than nucleic acid-based methods (17% vs 14%), in line with our understanding of p16 as a surrogate marker of HPV status, and suggesting that p16 may be a sensitive and specific marker for HPV+NPC. However, larger studies directly comparing p16 and direct HPV testing in HPV+NPC are needed.

We also sought to characterize clinical outcomes associated with HPV+NPC, such as overall survival (OS) and progression free survival (PFS). Unfortunately, amongst our included studies, we had very few studies that contributed this information, compounded by heterogeneous time intervals, precluding us from obtaining an adequately powered statistic. There have been a few North American studies that have investigated whether HPV+NPC and EBV+NPC differ in prognosis, though there is no clear consensus. In a US-based cohort of 343 NPC patients, of which 21 were HPV+NPC, there was no difference found in OS and DFS between HPV+NPC and EBV+NPC; a Canadian cohort with 451 NPC patients, of which 29 were HPV+NPC, also found no difference in outcomes between the two^11,50^. This is in contrast to a second US-based cohort of 61 NPC patients, of which 18 were HPV+NPC, where HPV+NPC was associated with decreased OS, PFS, and locoregional control compared to EBV+NPC^71^. The impact of viral status on clinical outcomes in NPC, especially in non-endemic areas, remains a subject of inquiry.

Our study possesses several limitations that warrant consideration. First, we identified heterogeneity in all of our analyses, a common challenge in meta-analyses. Our use of the random-effects model was intended to mitigate this issue; however, it is important to acknowledge that meta-analyses do not completely account for the inherent differences in study designs. The studies differed greatly in sample size as well as viral detection methodology, with the latter often limited to common high-risk HPV genotypes and precluding investigation of less common genotypes. We chose to include studies with small sample sizes due to the already limited total sample size for analysis, which could bias results. It is also worth noting that our study may exhibit a degree of geographic bias, as certain regions, such as Africa and Europe, were underrepresented. This is further compounded by having our studies limited to two languages; it is possible there are studies published in additional languages that could alter the findings demonstrated here.

In summary, our study provides a new reference point prevalence of HPV in NPC, stratified by geographic region, and describes notable epidemiological characteristics that provide an orthogonal argument for HPV as a significant etiological factor in NPC in North America.

## Data Availability

All data produced in the present work are contained in the manuscript.

## Ethics and Consent Statement

This study was determined to be exempt by the Mass General Brigham institutional review board.

## Data availability statement for the work

All data generated and analyzed during this study are included in this published article.

## Author Contributions

**Brian Y. Zhao**, conception, analysis, interpretation of data, drafting and revising manuscript, final approval, accountable for all aspects of the work; **Shun Hirayama**, conception, analysis, interpretation of data, drafting and revising manuscript, final approval, accountable for all aspects of the work; **Deborah Goss**, conception, drafting and revising manuscript, final approval, accountable for all aspects of the work; **Yan Zhao,** analysis, interpretation of data, drafting and revising manuscript, final approval, accountable for all aspects of the work; **Daniel L. Faden**, conception, analysis, interpretation of data, drafting and revising manuscript, final approval, accountable for all aspects of the work.

