## Supplemental Figure 1 for "Human Papillomavirus-Associated Nasopharyngeal Carcinoma: A Systematic Review and Meta-Analysis"

**Supplemental Figure 1.** Search terms used in the systematic review protocol.

**Embase 9.21.2022**

((epipharynx OR nasopharynx OR rhinopharynx OR epipharyngeal OR nasopharyngeal OR rhinopharyngeal OR 'postnasal space') NEAR/5 carcinoma*):ti,ab,kw OR 'nasopharynx carcinoma'/exp

**26,003**

'Papillomaviridae'/exp OR 'papillomavirus infection'/exp OR Papillomaviridae:ti,ab,kw OR 'human papillomavirus*':ti,ab,kw OR 'Human papilloma virus*':ti,ab,kw OR 'human papillomavirus-positive':ti,ab,kw OR 'Papillomavirus Infection*':ti,ab,kw OR HPV:ti,ab,kw OR HPV+:ti,ab,kw OR  'human wart virus*':ti,ab,kw OR alphapapillomavirus*:ti,ab,kw OR betapapillomavirus*:ti,ab,kw OR gammapapillomavirus*:ti,ab,kw OR mupapillomavirus*:ti,ab,kw OR 'human papilloma virus*':ti,ab,kw OR HPV-16:ti,ab,kw OR HPV-18:ti,ab,kw OR 'HPV-positive':ti,ab,kw OR ((wart* OR condyloma OR verruca) NEAR/5 virus*):ti,ab,kw

**112,042**

**1&2 = 582**

**Medline 9.21.2022**

exp Nasopharyngeal Neoplasms/ OR ((epipharynx OR nasopharynx OR rhinopharynx OR epipharyngeal OR nasopharyngeal OR rhinopharyngeal OR 'postnasal space') ADJ5 carcinoma*).ti,ab,kw.

**22925**

exp Papillomaviridae/ OR exp Papillomavirus Vaccines/ OR Papillomaviridae.ti,ab,kw. OR "human papillomavirus*".ti,ab,kw. OR "Human papilloma virus*".ti,ab,kw. OR "human papillomavirus-positive".ti,ab,kw. OR "Papillomavirus Infection*".ti,ab,kw. OR HPV.ti,ab,kw. OR HPV+.ti,ab,kw. OR "human wart virus*".ti,ab,kw. OR alphapapillomavirus*.ti,ab,kw. OR betapapillomavirus*.ti,ab,kw. OR gammapapillomavirus*.ti,ab,kw. OR mupapillomavirus*.ti,ab,kw. OR "human papilloma virus*".ti,ab,kw. OR HPV-16.ti,ab,kw. OR HPV-18.ti,ab,kw. OR "HPV-positive".ti,ab,kw. OR ((wart* OR condyloma OR verruca) ADJ5 virus*).ti,ab,kw.

**66498**

**1&2 = 238**

**Web of Science - All Databases 9.21.2022**

TS=(("Nasopharyngeal Neoplasm*" OR (epipharynx OR nasopharynx OR rhinopharynx OR epipharyngeal OR nasopharyngeal OR rhinopharyngeal OR 'postnasal space' NEAR/5 carcinoma*)))

**46,351**

TS=("papillomaviridae" OR "human papillomavirus*" OR "Human papilloma virus*" OR "human papillomavirus-positive" OR "Papillomavirus Infection*" OR HPV OR HPV+ OR "human wart virus*" OR alphapapillomavirus* OR betapapillomavirus* OR gammapapillomavirus* OR mupapillomavirus* OR "human papilloma virus*" OR HPV-16 OR HPV-18 OR "HPV-positive" OR ((wart* OR condyloma OR verruca) NEAR/5 virus*))

**76,847**

**1&2 = 747**

**Supplemental Figure 2.** (A) Forest plot of HPV+NPC prevalence in North American and UK studies using the random-effects model, stratified by race (B) Meta-regression model with race as a moderator; Asian identity was set as the intercept.

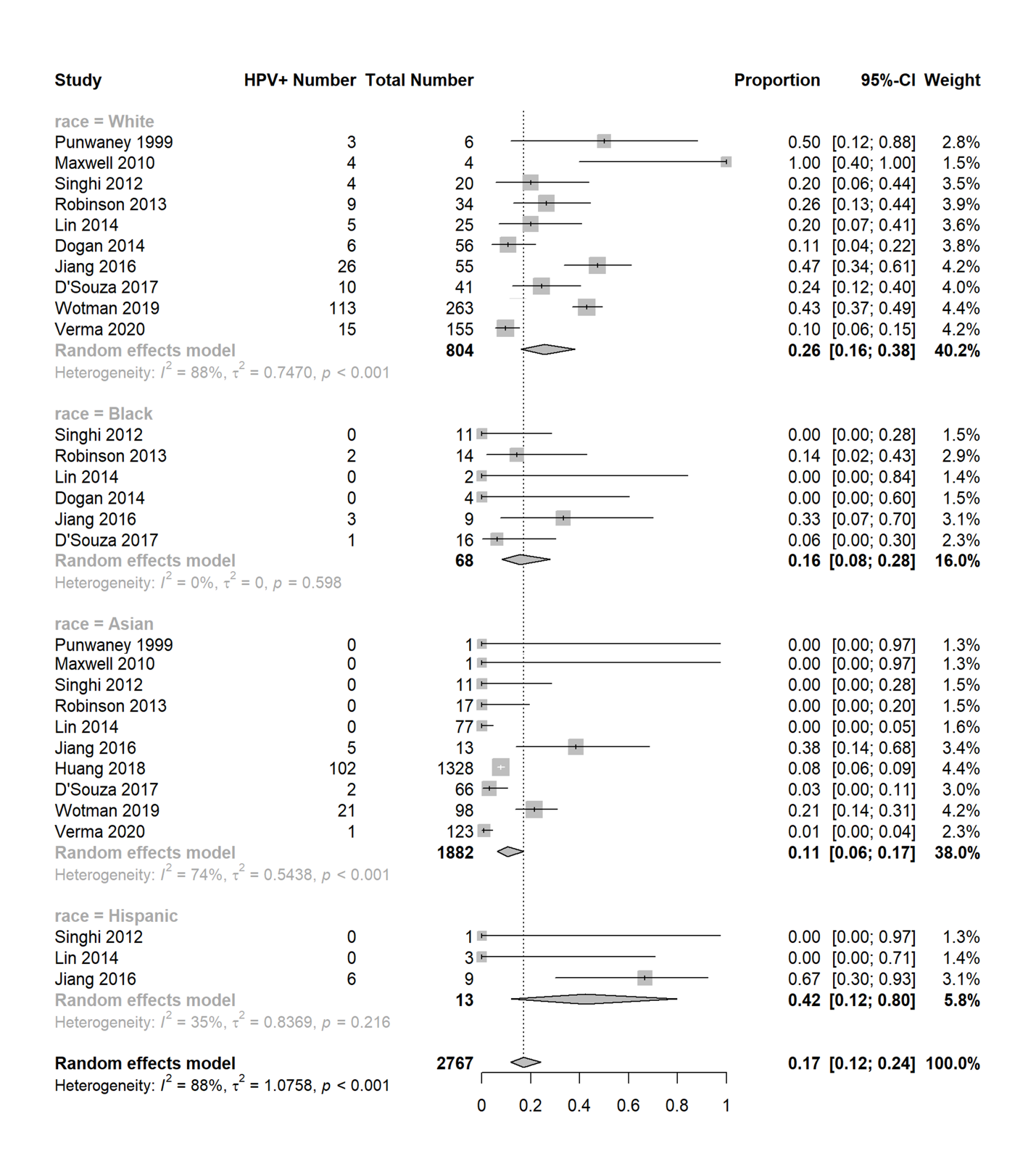

**A**

**B**

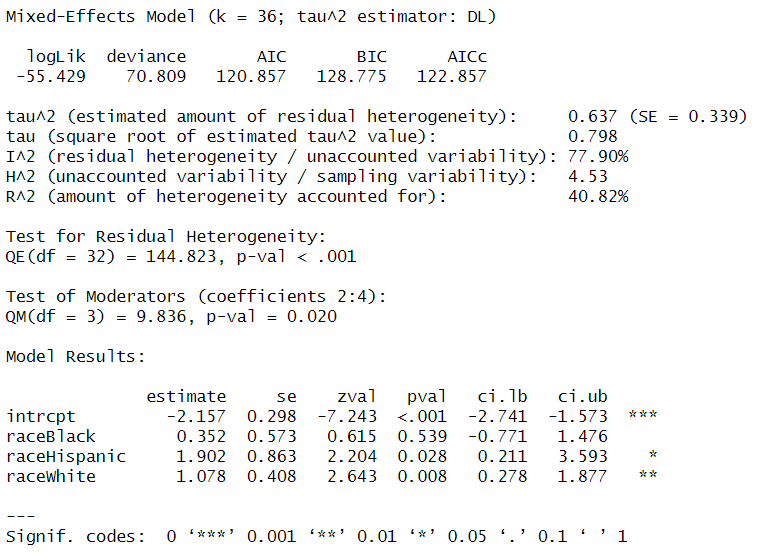

**Supplemental Figure 3.** (A) Forest plot of NPC+HPV prevalence using the random-effects model, stratified by reported sex (B) Meta-regression model with sex as a moderator; female identity was set as the intercept.

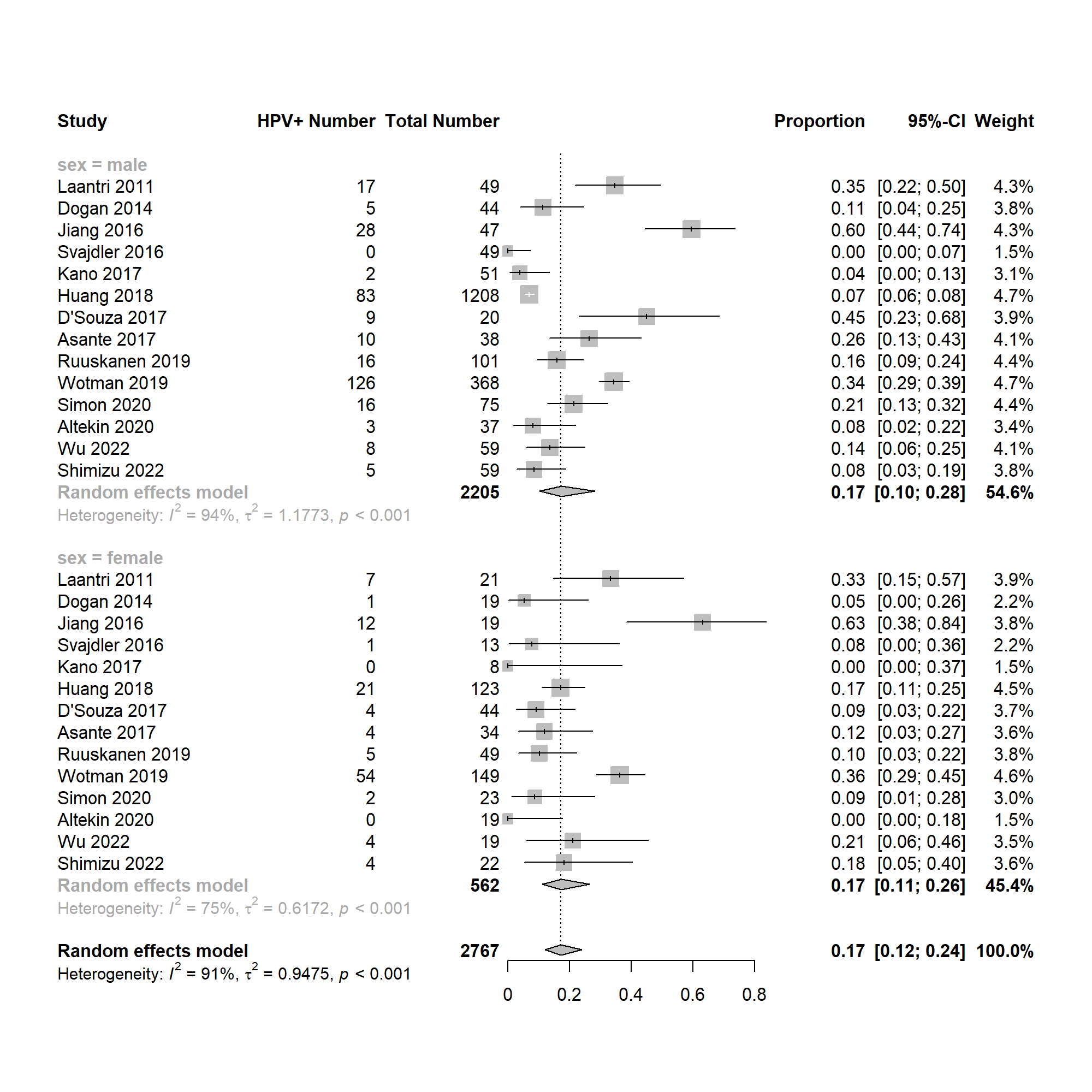

**A**

**B**

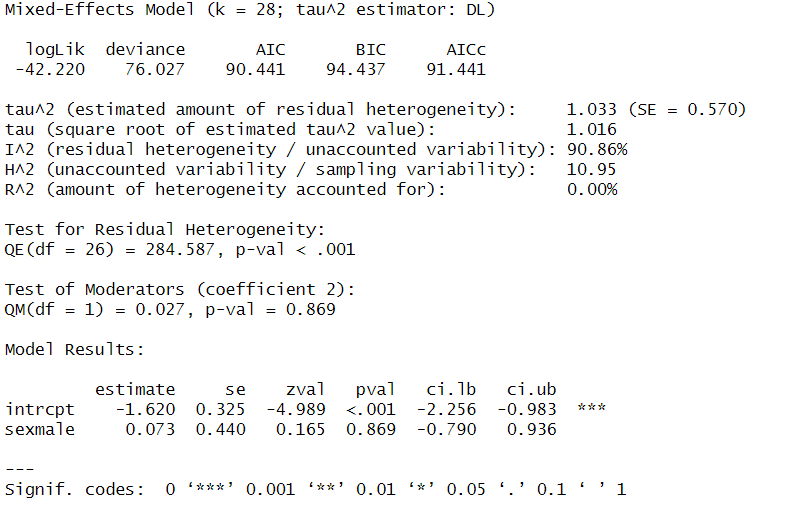

**Supplemental Figure 4:** (A) Forest plot of HPV+NPC prevalence using the random-effects model, stratified by WHO Tumor Classification (B) Meta-regression model with WHO Tumor Classification as a moderator; WHO I was set as the intercept.

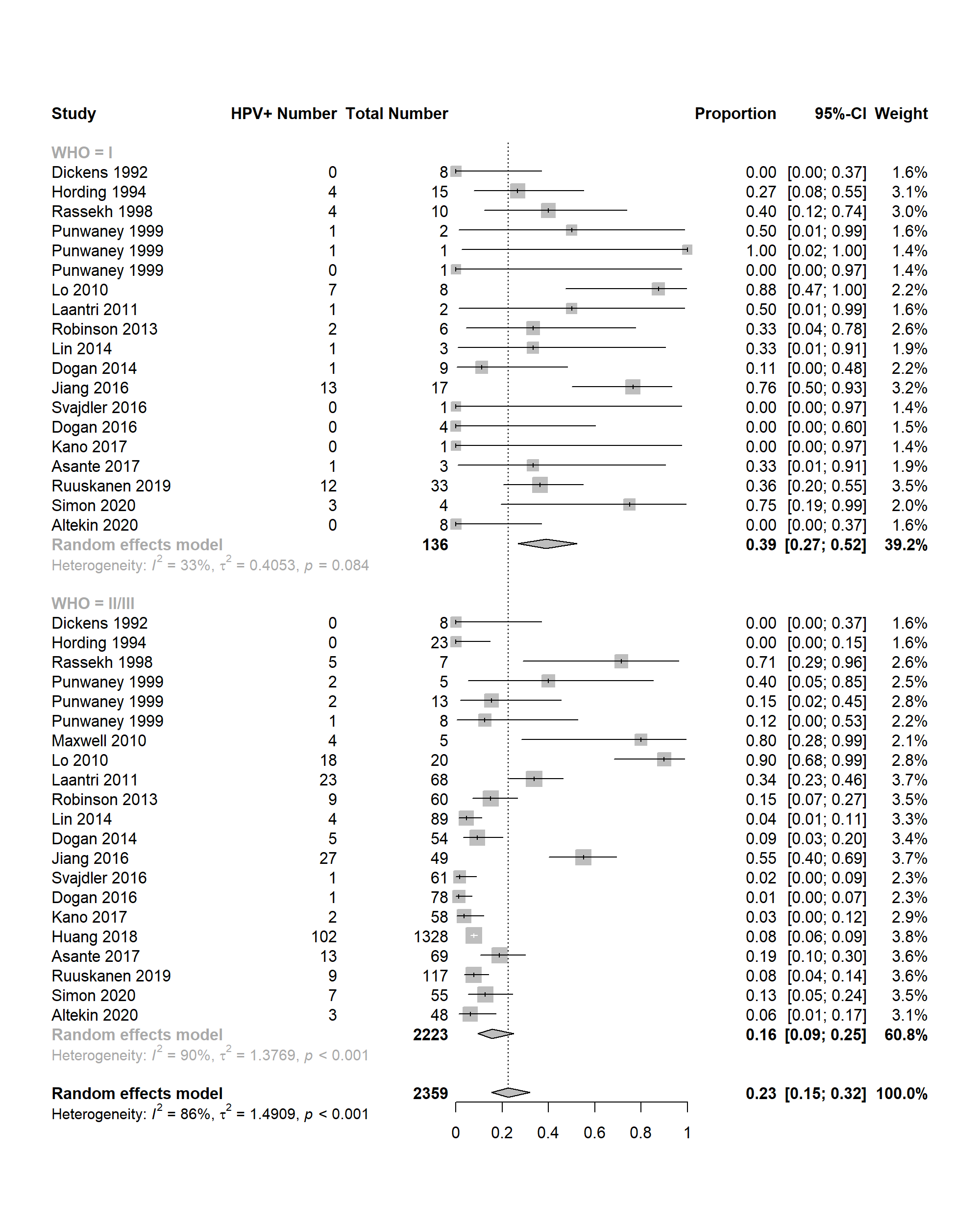

**A**

**B**

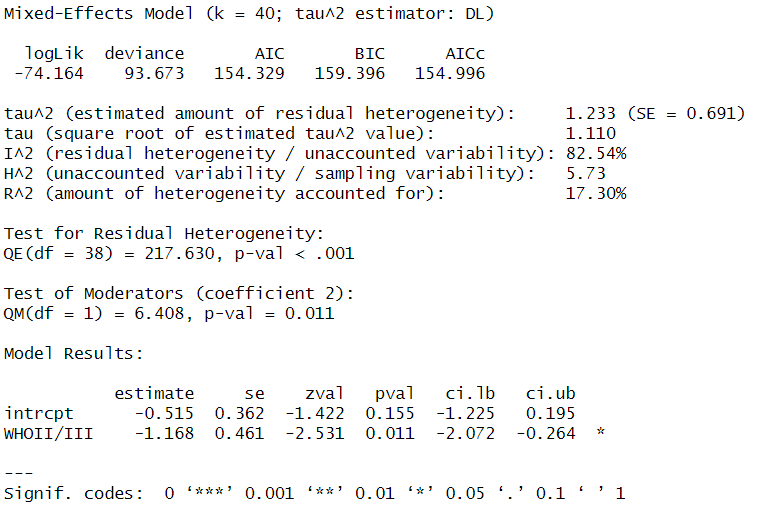

**Supplemental Figure 5.** (A) Forest plot of HPV+NPC prevalence using the random-effects model, stratified by HPV detection method (B) Meta-regression model with HPV detection method as a moderator; non-p16 methods was set as the intercept.

**A**

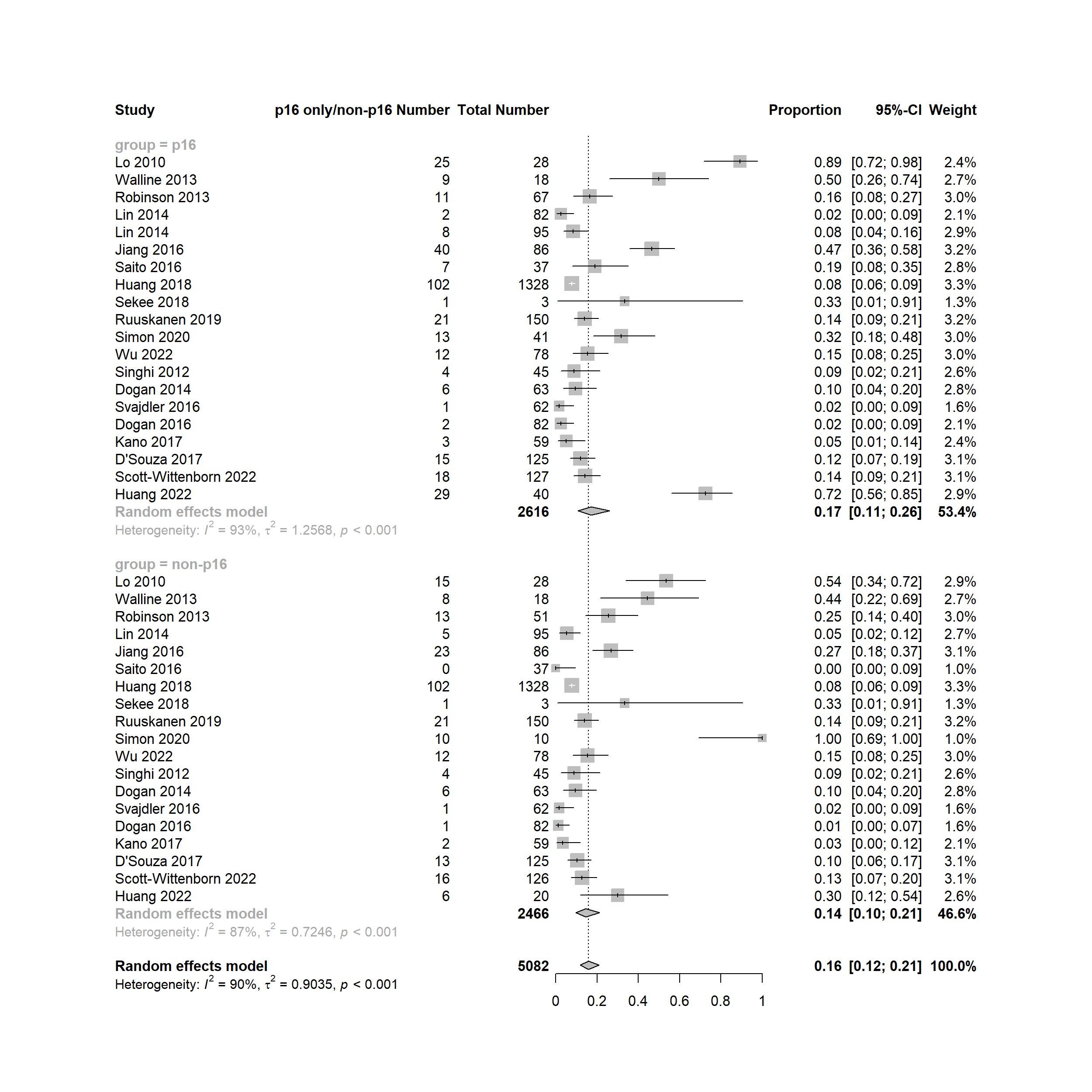

**B**

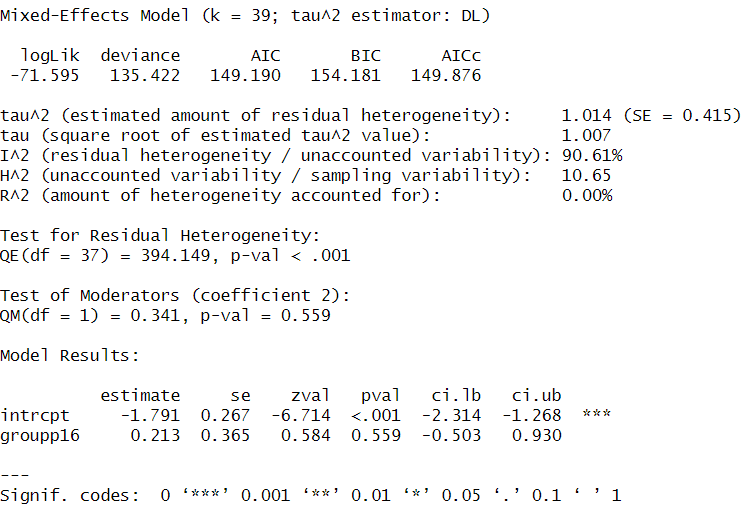

**Supplemental Figure 6.** (A) Forest plot of HPV+NPC prevalence using the random-effects model, stratified by presence of HPV only, EBV only, or HPV/EBV co-infection (B) Meta-regression model with viral presence as a moderator; EBV only was set as the intercept.

**A**

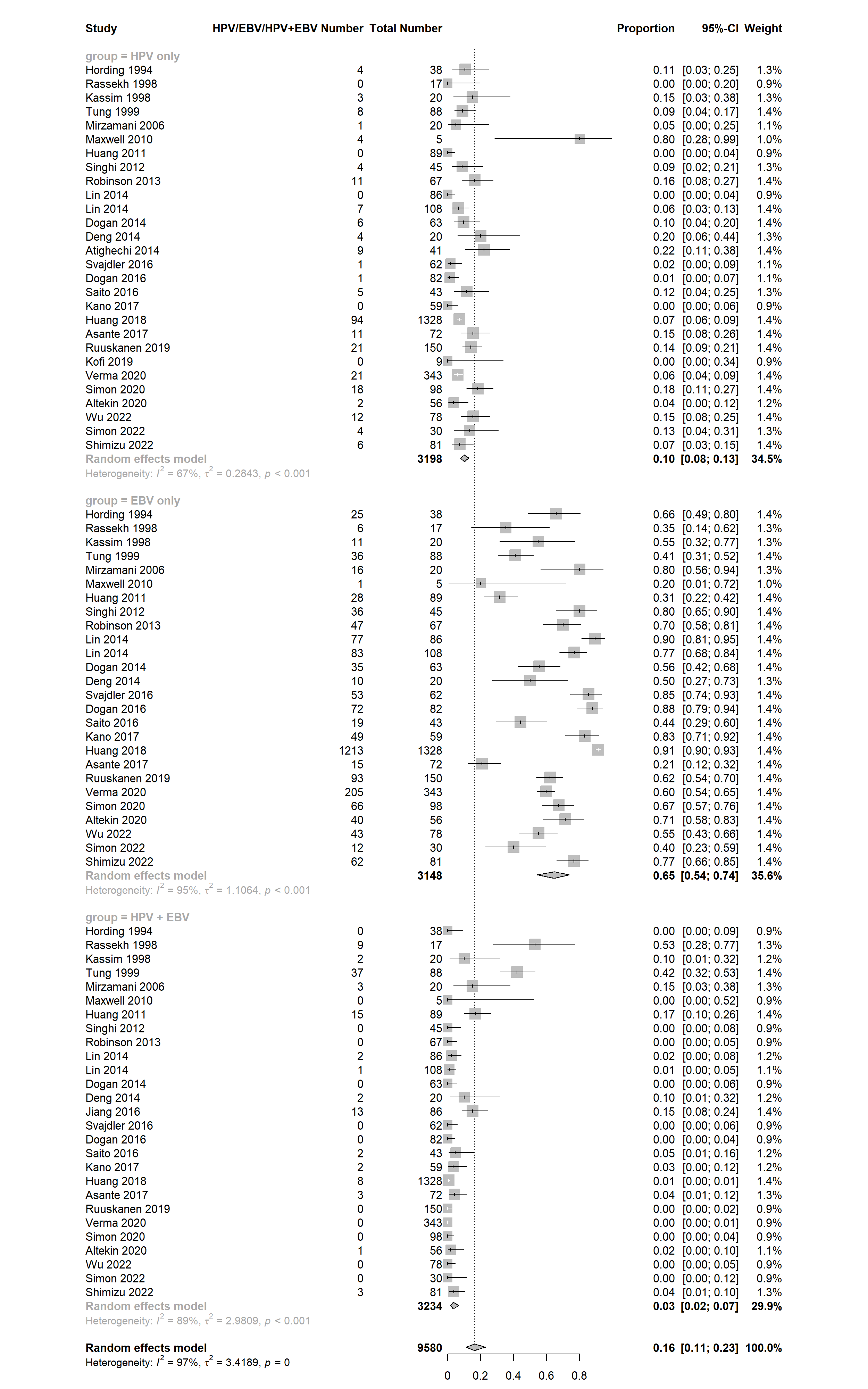

**B**

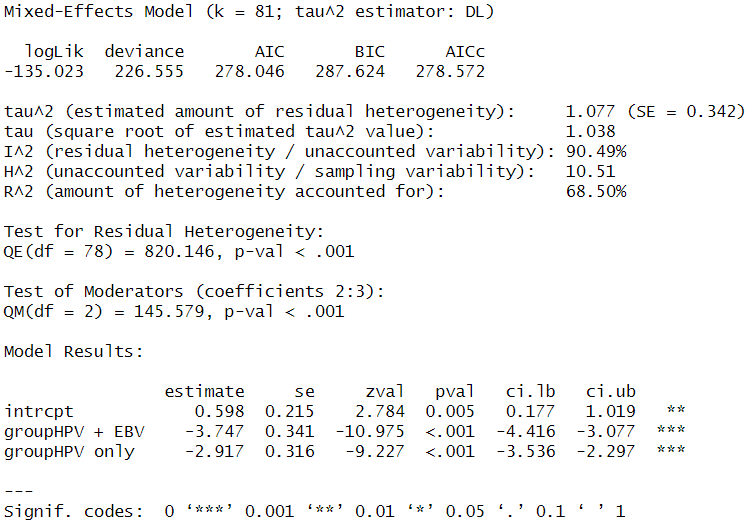

**Supplemental Figure 7.** (A) Funnel plot displaying the standard error by proportion for the 46 studies included in the analysis (B) Egger’s test results.

**A**

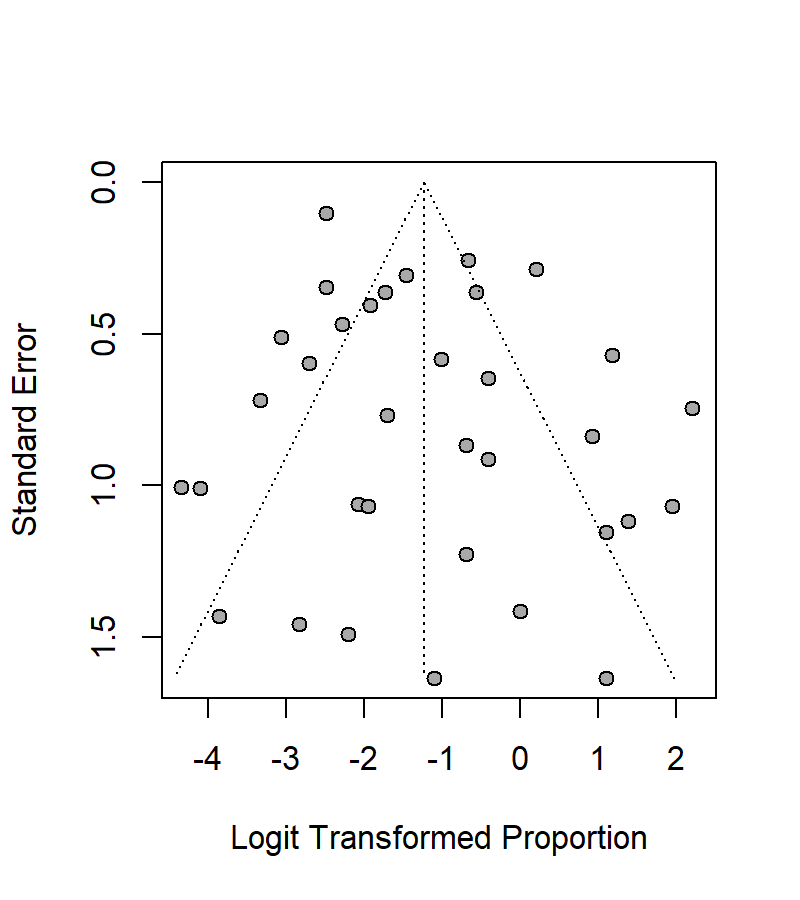

**B**

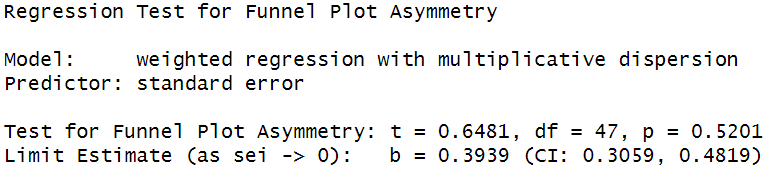
